# Short intervals of inertial measurement unit data collected during activities of daily living can provide a quantitative method to assess disease severity and functional ability in Parkinson’s disease

**DOI:** 10.64898/2026.10.30.26366941

**Authors:** Vignesh Vanchinathan, Matthew Farah, Arihant Singh, Chloe Zhang, Ramya Palani, Nana Osei-Owusu, Roma Desai, Jaya Hamkins, Daniella Iglesias Hernandez, Manuel Hernandez, Alexander Pantelyat, Stephen Grill

## Abstract

Parkinson’s disease (PD) is a progressive neurodegenerative disorder affecting roughly one million people in the U.S. People with PD (PwPD) exhibit classic motor symptoms that limit performance on functional tasks and reduce quality of life (QOL). Despite functional ability (FA) being a critical indicator of patient well-being, objective and quantitative measurement of FA during activities of daily living (ADLs) is not a part of standard clinical assessments. To address this gap, we conducted a pilot study of PwPD (n=39) and healthy controls (n=7) performing ADLs. We attached nine-axis inertial measurement units (IMUs) to toothbrushes and hair combs to collect continuous acceleration and angular velocity data during toothbrushing and hair-combing tasks. After preprocessing, we visualized signals with kernel density estimation and continuous wavelet transforms, revealing characteristic distributional, temporal, and spectral patterns. We then segmented signals into 3-9 second non-overlapping intervals. From each interval, we extracted 30 features (13 acceleration-derived, 17 angular-velocity-derived) and trained eXtreme Gradient Boosted (XGBoost) classifiers to discriminate disease state and severity, symptom presence, and impaired FA. Notably, models trained on hair-combing data discriminated PwPD from controls (balanced accuracy 0.864) and identified patient-reported impairment in FA (0.869). Models trained on toothbrushing data achieved moderate performance on the same tasks (balanced accuracy 0.752 and 0.686, respectively). These preliminary findings suggest that short-segment IMU data from common ADL implements can quantitatively characterize disease severity and task-specific performance in PwPD, supporting their integration into routine clinical care.

## I. Introduction

Parkinson’s disease (PD) is the second most common neurodegenerative disorder after Alzheimer disease, affecting approximately one million people in the United States and 11.8 million globally [1,2]. PD results from the progressive loss of dopaminergic neurons in the substantia nigra pars compacta (SNpc), a region of the basal ganglia essential for the execution of smooth, coordinated movements. Consequently, people with PD (PwPD) develop motor symptoms such as bradykinesia, rigidity, and tremor [3]. Although pharmacotherapy can alleviate most motor symptoms, PD remains incurable, and many patients experience progressive declines in functional ability (FA) and daily autonomy [4, 5].

The ability to perform activities of daily living (ADLs) without assistance is a critical determinant of quality of life (QOL) in PD. Loss of FA increases comorbidity risk and is closely associated with depression, social isolation, and reduced dignity [6-8]. Occupational therapy has emerged as an evidence-based intervention that can limit FA decline in PD, with therapists providing personalized strategies to help patients maintain mobility and ADL performance despite motor impairments [9-11].

Widely used clinical rating scales, including the Unified Parkinson’s Disease Rating Scale (UPDRS) and its Movement Disorder Society-sponsored revision (MDS-UPDRS), incorporate ADL and QOL assessments. However, these scales are nonlinear, discontinuous, and often rely on subjective patient reporting, which introduces bias and limits measurement precision [12, 13]. Time and operational constraints further lead clinicians to prioritize motor examinations (e.g., UPDRS Part III) during revisits, often neglecting direct evaluation of FA despite its importance [14]. Motor examinations are also subject to intra-rater variability. Consequently, current assessments remain non-standardized, qualitative, and limited in interpretability and clinical actionability. This gap highlights the need for objective, quantitative tools to monitor FA during ADLs, which serve as proximate measures for overall independence [15]. Such tools could provide a consistent and quantitative way to measure patient outcomes across therapeutic interventions and inform the caretaking team.

Sensor-based technologies have emerged as a promising solution to this unmet need, offering noninvasive, quantitative, and continuous real-time motion measurement [16-18]. Inertial measurement units (IMUs)—which capture triaxial acceleration, angular velocity, and orientation data—have commonly been mounted on the limbs or trunk to measure movement for a low-cost [19]. Using signal processing and machine learning (ML) techniques, IMU data have been used to identify cardinal PD symptoms, distinguish PwPDs from healthy controls, and quantify longitudinal changes in motor state [20-23].

Critically, few comprehensive datasets capture PwPD performing ADLs, and consequently, few models can estimate FA using movement features derived from these QOL-relevant tasks [24, 25]. Such data would provide direct insight into patient QOL. Although some IMU datasets exist for specific ADLs (e.g., toothbrushing), they are often limited to healthy populations, precluding comparison with PwPD [26, 27]. Other studies do include PD participants but are restricted by small sample sizes or narrow clinical scope [28, 49, 50].

To address this, we collected movement data from PwPD and controls during two ADLs— brushing teeth and combing hair—generating a larger dataset than previously reported for these tasks. We characterized distributional differences; extracted temporal, statistical, and spectral features; and used them to identify those with impaired task performance. Our preliminary results suggest that impaired task performance and disease severity can be effectively quantified using short segments of this modality, which may support scalable, objective assessment of FA in home-based studies.

## II. Methodology

### A. Study Design

PD patients (n = 39) and controls (n = 7) capable of performing ADLs were analyzed in this report, which includes participants enrolled through March 19^th^, 2026. This cohort represents the initial cohort collected by investigators; enrollment is ongoing. Participants were recruited from multiple sites within the Johns Hopkins Medicine system under IRB-approved protocol #00442448, with additional controls recruited from the Johns Hopkins University Homewood Campus. All participants provided written informed consent. Exclusion criteria included diagnoses of other neurological or psychological conditions affecting movement or participation ability. Controls—typically care partners of enrolled patients to ensure age-matching—were defined as individuals without clinically diagnosed PD or other movement disorders.

Prior to task performance, Parts I-III of the UPDRS were administered to PwPD to assess symptom severity [29]. Clinical notes regarding motor patterns, brushing technique, dominant hand, hair type, and at-home toothbrush type (manual/electric) were documented for contextual reference. Controls were not evaluated by a neurologist and were assigned naive UPDRS subscores of 0. No protected health information (e.g., age, name, sex) was collected; however, trials were videotaped to provide a visual reference for the recorded motion data.

UPDRS Parts I-III subscores were summed to compute several composite scores reflecting distinct clinical domains: rigidity (RG), bradykinesia (BK), functional ADLs (FT), resting tremor (TR), action/postural tremor (AT), Part III total (P3) and total Part I-III UPDRS score (TOT) **(Table 1).** For example, subscores “3.23 Finger Taps”, “3.24 Hand Movements”, and “3.25 Rapid Alternating Movements of Hand” for the corresponding side used during the task were used to calculate the BK composite score. For upper body related scores, only subscores corresponding to the side used during task performance were included (**Supplemental Table 1**).

**Table 1.** Participant cohort characteristics for 39 PwPD and 7 controls. Classes 0-3 were delineated using total Part I-III UPDRS score (TOT). Participants are considered positive (+) if their composite score for that category is positive. Controls were naively assigned 0 for all UPDRS subscores because they were not evaluated by a neurologist.

| **Characteristics** | **PwPD** | **Controls** |
|---|---|---|
| Right-Handed | 35 (89.7 %) | 7 (100.0 %) |
| Left-Handed | 4 (11.4 %) | 0 (0.0 %) |
| Class 0 (TOT: 0) | 0 (0.00%) | 7 (100.00%) |
| Class 1 (TOT: 1-32) | 30 (76.9 %) | 0 (0.0 %) |
| Class 2 (TOT: 33-56) | 8 (20.5 %) | 0 (0.0 %) |
| Class 3 (TOT: 57+) | 1 (2.6 %) | 0 (0.0 %) |
| Rigidity/RG (+) | 29 (74.4 %) | 0 (0.0%) |
| Rest Tremor/TR (+) | 14 (35.9 %) | 0 (0.0%) |
| Action Tremor/AT (+) | 10 (25.6 %) | 0 (0.0%) |
| Bradykinesia/BK (+) | 28 (71.8 %) | 0 (0.0%) |
| Functional Impairment/FI (+) | 34 (87.2 %) | 0 (0.0%) |

Motion data (i.e., acceleration and angular-velocity data) was collected using a WitMotion WT9011DCL Bluetooth 5.0 Accelerometer, a 9-axis, off-the-shelf inertial measurement unit (IMU) (manufactured by WitMotion Shenzhen Co., Ltd, ShenZhen China) containing an accelerometer, gyroscope, magnetometer, and other sensors [30]. Accelerometers were connected to mobile devices via the WitMotion application, and they were calibrated before each data collection session using a built-in “acceleration calibration” setting. Sensor “bandwidth” (sampling frequency) was set to 98 Hz. The sensor was attached to a standard toothbrush or standard comb with a 3D-printed plastic holder. Separate holders were designed to be kept secure on the toothbrush and comb with friction. The sensor was held on the holder with a rubber band near the bottom of the shaft. The position of the sensor partially obstructed the handle but largely did not affect participant arm motion as they performed the activities. **(Fig. 1a, 1b).**

**Figure 1.**
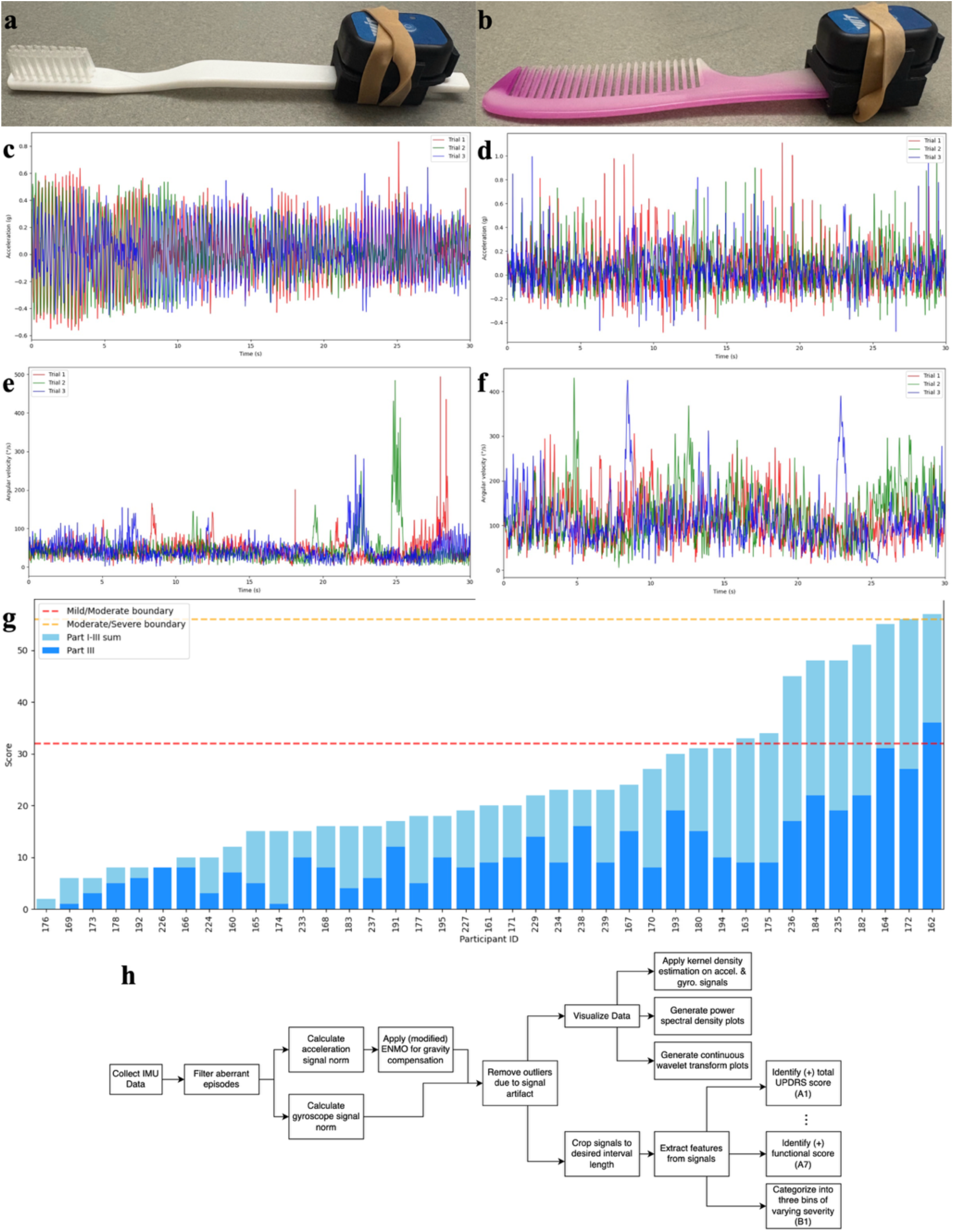
Sensor setup, exemplar movement data, and clinical cohort overview. Setup of instrumented ADL tools: an inertial measurement unit (IMU) mounted on (a) a toothbrush and (b) a hair comb via a custom 3D-printed mount. (c—f) Representative raw time-series data of acceleration magnitude (top) and angular velocity magnitude (bottom) captured during (c, e) brushing and (d, f) combing tasks. (g) Distribution of overall disease severity in the PwPD cohort, assessed by the summed UPDRS Parts I-III total score. Dashed red and yellow lines mark the boundaries between mild and moderate (score: 32) and moderate and severe (score: 56) impairment, respectively. (h) Graphical illustration of computational analyses employed throughout the paper, including preprocessing, data visualization, and machine learning (ML) analyses.

First, participants performed either toothbrushing or hair-combing without a specified duration to approximate typical home performance times **(Fig. 1c-f)**. Following this initial trial, participants completed two or three additional one-minute trials, with the number determined by the length of the first trial. If the participant’s first trial was already longer than one-minute, only two additional trials were performed; if not, three were performed. Continuous motion data were captured throughout all trials. After data ingestion, they were preprocessed, visualized, and used for a machine learning inference pipeline **(Fig. 1h).**

### B. Preprocessing & Feature Extraction

Signals shorter than 30 seconds were excluded to remove any trials likely to be incomplete or not focused on the ADL for an extended period. Those with sensor artifacts that rendered them unusable were excluded as well (n = 6 participants). For both angular-velocity and acceleration time-series, the x-, y-, and z-components were combined into a single magnitude signal. Gravity’s contribution was removed from the acceleration magnitude using the Euclidean Norm Minus One (ENMO) method [31]. However, negative values were not clipped to zero to preserve a symmetric distribution about zero. Outliers— defined as absolute values exceeding three standard deviations from the mean magnitude—were replaced with the mean. Signals were segmented into non-overlapping intervals of 3, 5, 7, and 9 seconds [32, 33]. From each window, thirty features were extracted, and a separate machine learning analysis was performed for each interval length (**Supplemental Table 2**).

### C. Kernel density estimation

Both the acceleration and angular velocity signals were used to construct kernel density estimator (KDE) plots using a Gaussian kernel with Silverman’s rule [34]. The KDE plots were used to both visualize the distributions of signal values, generate statistical features, and check for statistically significant differences. KDE plots were generated for both brushing and combing activities separately to account for task-specific variation. These KDE plots were also used to qualitatively compare acceleration and angular velocity distributions between experimental groups, notably between PD patients and controls and between different disease severities. Unlike other analyses, signals for KDE visualization were not preprocessed with ENMO. Instead, the mean was subtracted from each signal to center its KDE at 0 g. This approach facilitated direct comparison of distributional spread across populations and protected against outlier signals that were not centered at 0.

### D. Power spectral density

To assess signal features in the frequency domain, power spectral density (PSD) plots were generated from the acceleration magnitude signal using a Welch’s method-inspired approach [35]. A Hanning window was applied to each full-length signal to reduce spectral leakage, with appropriate correction for the window’s energy, followed by computation of the Fast Fourier Transform (FFT). A one-sided PSD was derived by taking the squared magnitude of the FFT coefficients at positive frequencies, applying a scaling factor of 2 excluding the DC and Nyquist components. Finally, PSD values were normalized by the window energy, sampling rate, and number of points to yield a proper power spectral density estimate.

### E. Continuous wavelet transform

To assess how frequency contribution evolved during task performance, continuous wavelet transform (CWT) plots were generated based on the first 30 seconds of each signal; signals with less than 30 seconds of data were excluded [36]. To generate the CWT, a complex Morlet wavelet (‘cmor1.5-1.0’) was used as the mother wavelet. The analysis was performed over a frequency range of 0.5 to 20 Hz, relevant for human movement, using 80 linearly spaced frequency points. For each time point *t*, the squared magnitude of the CWT coefficients across all frequencies was computed to obtain a time-varying power spectrum. The spectrum at each time point *t* was normalized to integrate to 1 across frequencies, converting it into a probability density function (PDF). Therefore, these plots represent the relative distribution of signal power across frequencies at each moment in time, independent of the total power.

### F. Computation of complex plot CWT ellipse area

In addition to analyzing the entire time-frequency matrix generated by CWT analysis, a single compact feature (the ellipse area of the CWT complex plot) was computed from each signal’s CWT representation. The CWT produces a complex signal containing a real component (*R*(*n*)) and an imaginary component (*IM*(*n*)), representing the magnitude and phase of wavelet distribution, respectively. The complex plot is generated by plotting *R*(*n*) against *IM*(*n*). It has been previously shown that the complex plot representations of the CWT form an ellipse, and the area of these ellipses can valuable insight into disease severity. To compute the area of these ellipses, the following procedure was used [37].

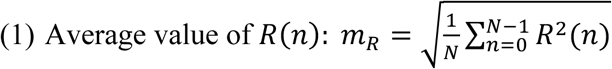

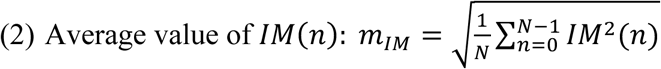

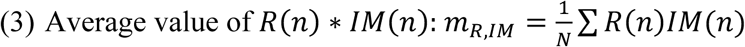

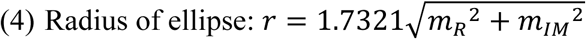

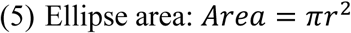

### G. XGBoost ensemble model inference task

We framed the analysis as both a binary and multiclass classification task. The vector of features extracted from the sensor data was the model input, and a binary prediction was output for each composite score: whether the score was zero or positive. Labels were assigned at the participant level. For higher-resolution classification, the TOT score was used to group participants into four classes (Class 0: TOT 0; Class 1: TOT 1-32; Class 2: TOT 33-56; Class 3: TOT 57+). All analyses were performed separately for the toothbrushing and hair combing tasks. Furthermore, to determine the optimal signal segmentation, the entire modeling pipeline was independently executed for non-overlapping window lengths of 3, 5, 7, and 9 seconds, with the goal of identifying the interval length that delivered the best overall performance across all tasks (**Supplemental Table 3**).

#### 1) Classification

We use gradient-boosted decision trees as implemented in the open-source XGBoost package [38] with default hyperparameters; no tuning was performed. Data were split 80/20 into training and test sets and were grouped by participant so that no participant appeared in both. This split was not stratified by class. Features were standardized using statistics from only the training set. An XGBoost model with scaled positive- and negative-class weight (for binary tasks; class weighting was not applied in the multiclass task) was trained and then tested on the held-out set. We report balanced accuracy (BA; unweighted mean of per-class recall) due to class imbalance **(Fig. 1g).** We then performed stratified, participant-grouped, tenfold cross-validation. Cross-validation was repeated ten times with different seeds to yield 100 scores per model. We choose to report the average and standard deviation of the BA across the cross-validation as a robust and stable measure of performance across different folds. Ninety-five percent confidence intervals were constructed using the mean BA for each repeat, and paired t-tests (with Holm-Bonferroni p-value correction) were used to assess significant differences between the grand mean of each interval length. Extreme outliers were 95% winsorized.

#### 2) Feature Importance

To improve explainability of the XGBoost models, we employed the Shapley Additive Explanations (SHAP) approach and its corresponding open-source package [39, 40]. For each model trained, mean absolute SHAP values were calculated on the held-out test set and used to determine the ten most relevant features for the classification task. These were displayed on beeswarm plots for visual inspection. Volcano plots were also produced to investigate significance versus effect size of all features between PwPD and controls.

### H. Software

All analysis was conducted using Python 3 and standard packages such as NumPy, Pandas, Scikit-Learn, XGBoost, and SHAP. Continuous wavelet transformations were applied to the signals using the PyWavelets package [41]. Visualization was done through Matplotlib and Seaborn. Analysis was conducted using the SAFE Desktop virtual machine system provided by Johns Hopkins Research Information Technology (Windows 11, 6 CPUs, 24GB RAM, 1GB GPU).

## III. Experimental Results

### Kernel density estimation reveals progressive, bradykinesia-sensitive distributional differences in ADL-based movement data

PwPD classically present with bradykinesia, the presence of slowed or diminished movements [42]. We hypothesized that bradykinesia would be evident in distributional changes in accelerometry (measuring acceleration) and gyroscope (measuring angular velocity) data. To assess distributional changes between PD patients and controls, and patients of varying disease severity, we generated kernel density estimation (KDE) plots of acceleration or angular velocity data. KDE analysis smooths discrete time-series data into a continuous probability density function (PDF) by summing Gaussian kernels— each centered on an individual data point—and normalizing the result.

We predicted that patients with more severe bradykinesia would display a narrower, more peaked acceleration distribution centered near 0 g, reflecting fewer extreme changes in velocity. While brushing teeth, PwPD (*σ* = 0.2941) exhibited a slightly diminished acceleration distribution compared to controls (*σ* = 0.3164), consistent with more severe bradykinesia (p = 0.36) (**Fig. 2a**). These distributional differences were further pronounced during combing trials; PwPD (*σ* = 0.2751) exhibited nearly half the variation in acceleration compared to controls (*σ* = 0.4792; p = 0.001) (**Fig. 2b**). Closer examination revealed that brushing trials from two participants exhibited modes that deviated from their mean at 0 g, likely inflating the group-level variance and thereby attenuating distributional differences between PwPD and controls during brushing. These outlier patients, both of moderate severity, likely also contributed to moderate PwPD (*σ* = 0.3363) exhibiting a slightly higher standard deviation in acceleration during brushing trials compared to mild PwPD (*σ* = 0.2835; p = 0.723). This interpretation is supported by visual analysis of the KDE plots; while the moderate PwPD distribution exhibits a sharper peak at 0 g and decays more rapidly than mild PwPD in the positive x-direction, it also display a small bump inconsistent with a unimodal Gaussian distribution in the negative x-direction consistent with these outlier patients, likely inflating the calculated standard deviation (**Fig. 2c**). Similarly, during hair combing trials, moderate PwPD (*σ* = 0.1980) displayed a less diffuse distribution than mild PwPD (*σ* = 0.2949), supporting the severity-dependent narrowing of movement dynamics (p= 0.053) (**Fig. 2d**).

**Figure 2.**
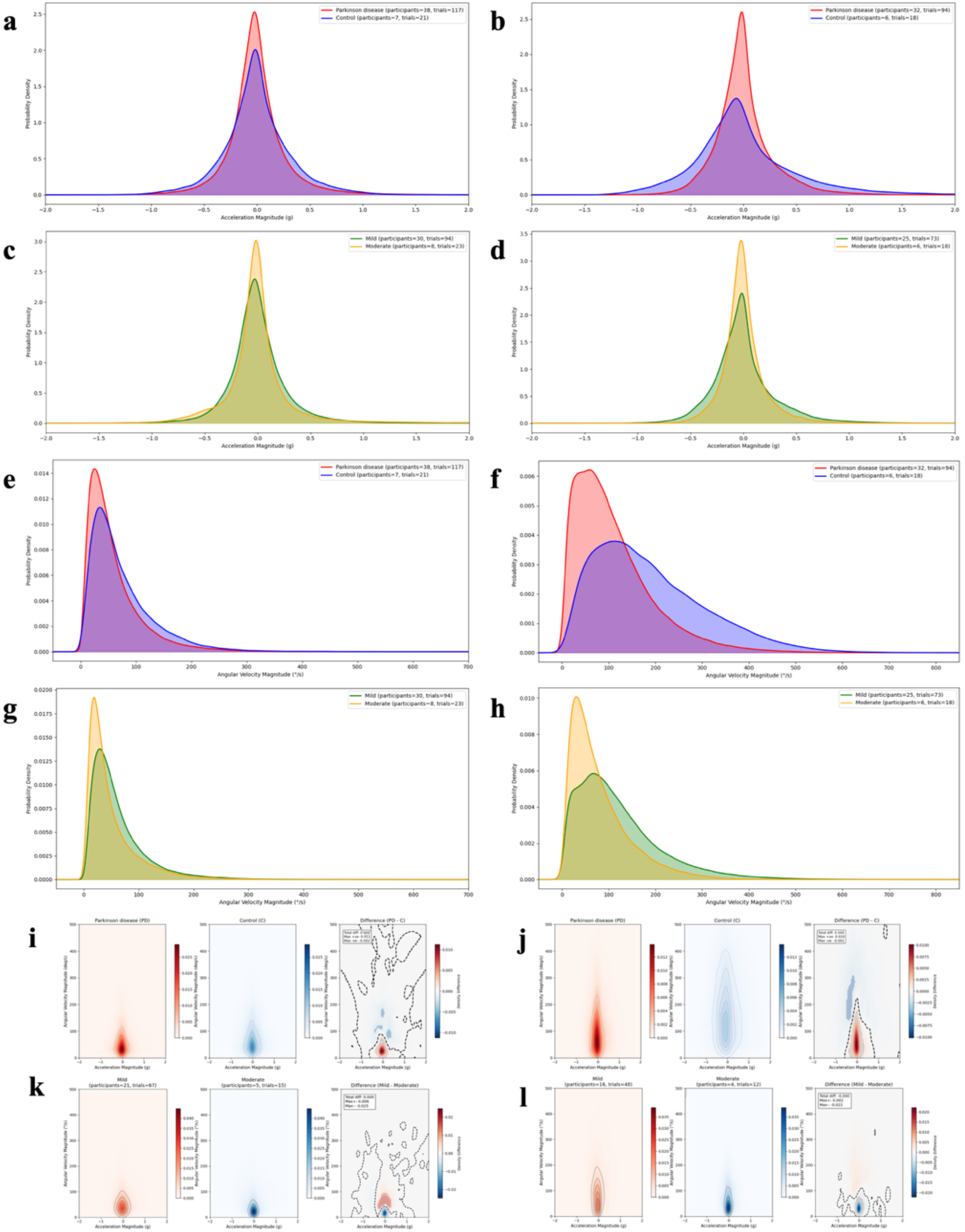
Kernel density estimation (KDE) reveals distributional differences in sensor data between PwPD of varying severity and controls during ADLs. Plots are based on data collected during two tasks: brushing teeth (left panels) and combing hair (right panels). (a, b) Two-dimensional (2D) KDE of acceleration data for PwPD vs. controls. (c, d) 2D KDE of acceleration data for PwPD stratified by disease severity. (e, f) 2D KDE of angular velocity data for PwPD vs. controls. (g, h) 2D KDE of angular velocity data for PwPD stratified by disease severity. (i, j) Three-dimension (3D) KDE combining acceleration and angular velocity for PwPD vs. controls. (k, l) 3D KDE combining acceleration and angular velocity for PwPD stratified by disease severity.

We similarly anticipated that patients with more severe bradykinesia would exhibit heavily right-skewed angular velocity distributions, with high probability density near 0 °/s. This pattern was observed during tooth brushing, where the angular velocity distribution of PwPD (*μ* = 50.1246; *M* = 45.2665; *σ* = 51.8051) was shifted toward lower values and showed similar variance compared to controls (*μ* = 74.5968; *M* = 57.7341; *σ* = 59.0780) (*p_μ_* = 0.077, *p_M_* = 0.068, *p*_σ_ = 0.142) (**Fig. 2e**). Consistent with increasing bradykinesia, moderate PwPD (*μ* = 49.3942; *M* = 34.0756; *σ* = 45.2559) had an even more left-shifted and less variable distribution than mild PwPD (*μ* = 62.6003; *M* = 47.7953; *σ* = 52.8937) (**Fig. 2g**). During hair combing, this effect was equally pronounced: PwPD distributions (*μ* = 110.8401; *M* = 90.5921; *σ* = 85.0817) were markedly left-shifted and less variable relative to controls (*μ* = 187.7073; *M* = 164.6107; *σ* = 118.0935) (*p_μ_* = 0.007, *p_M_* = 0.024, *p*_σ_ < 0.001) (**Fig. 2f**). Again, moderate PwPD (*μ* = 80.8019; *M* = 62.3939; *σ* = 63.7316) displayed a more left-shifted and narrower distribution than mild PwPD (*μ* = 119.6160; *M* = 99.0159; *σ* = 89.6267) (**Fig. 2h**). Together, these findings reflect a severity-dependent narrowing of rotational movement in patients with bradykinesia.

To further validate and synthesize these results, we generated three-dimensional (3D) KDE plots, mapping acceleration magnitude on the *x-*axis, angular velocity on the *y-*axis, and probability density as a heatmap (**Fig. 2i—l**). Consistent with our earlier findings, PwPD with more severe bradykinesia exhibited tighter, less diffuse probability distributions, with the highest density concentrated near the origin (0 g, 0 °/s).

### Power spectral density analysis uncovers energy-independent movement frequency differences during ADL performance

Given the observed distributional differences in acceleration between PwPD and controls, we expected corresponding differences in acceleration signal energy. During brushing tasks, PwPD exhibited lower and less variable accelerometry signal energy (*M* = 2.6896 ∗ 10^2^; *Q*1 = 1.3810 ∗ 10^2^; *Q*3 = 5.6444 ∗ 10^2^) compared to controls (*M* = 6.8953 ∗ 10^2^; *Q*1 = 2.3004 ∗ 10^2^; *Q*3 = 1.2968 ∗ 10^3^) (**Fig. 3a**). This difference was equally evident during combing trials, where PwPD again showed reduced and less variable signal energy (*M* = 2.3968 ∗ 10^2^; *Q*1 = 1.3976 ∗ 10^2^; *Q*3 = 3.9467 ∗ 10^2^) relative to controls (*M* = 1.0316 ∗ 10^3^; *Q*1 = 7.8699 ∗ 10^2^; *Q*3 = 1.8202 ∗ 10^3^) (**Fig. 3b**).

**Figure 3.**
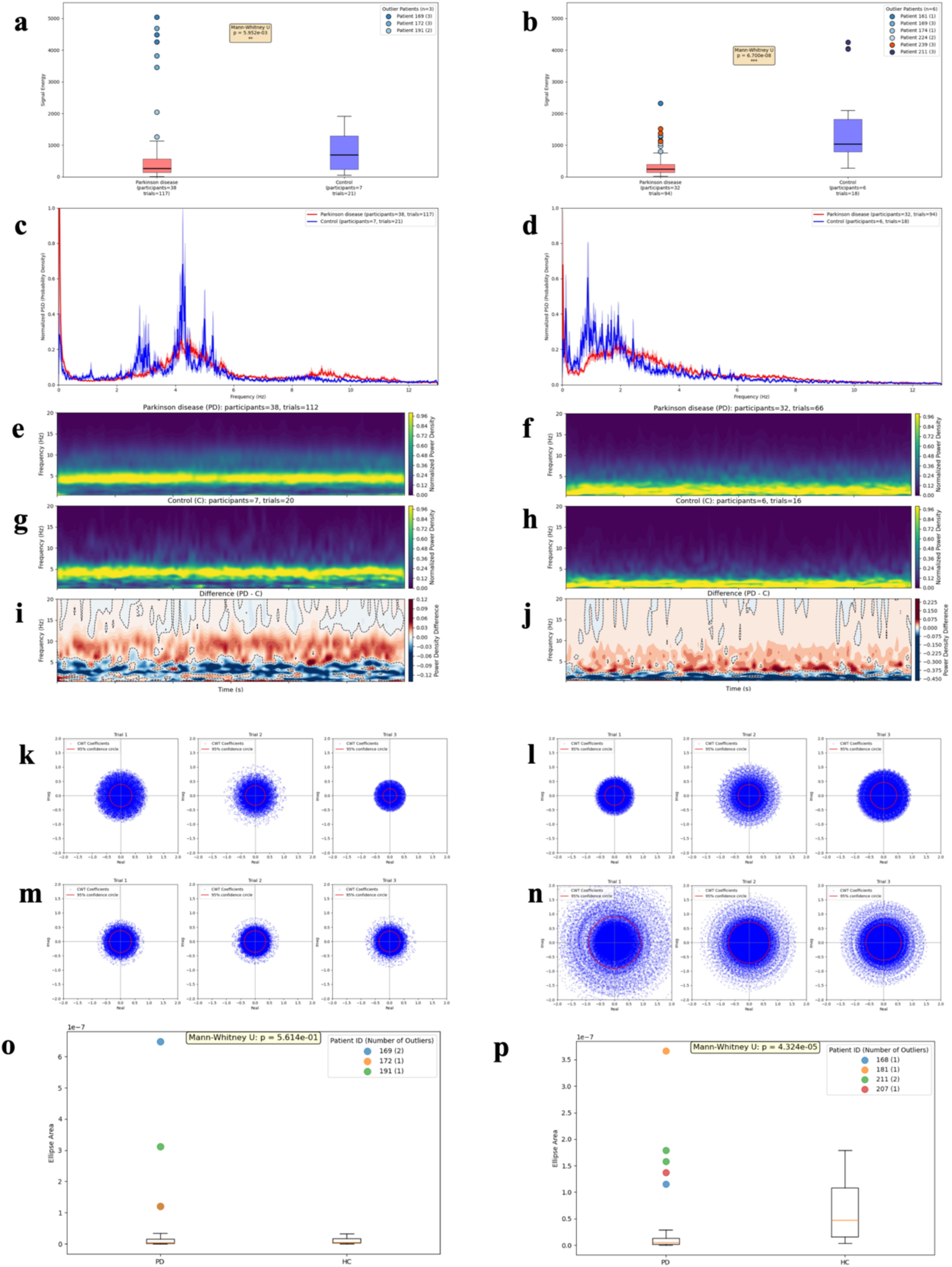
Frequency domain analyses of acceleration data during ADLs discriminates between PwPD and healthy controls. Data from two tasks—brushing teeth (left column) and combing hair (right column)—are analyzed. (a, b) Boxplots of total signal energy. Outliers are marked as beyond mean ± standard deviation. (c, d) Energy-normalized PSD plots. (e-h) Normalized CWT scalograms for PwPD (e, f) and controls (g, h). (i, j) Difference in frequency contribution (PwPD vs. controls) derived from CWT. (k-n) CWT complex area plots for PwPD (k, l) and controls (m, n) (o, p) Boxplots of the CWT complex plot area metric.

To compare movement frequencies between groups, we computed power spectral density (PSD) normalized to signal energy over physiologically relevant frequencies (0-20 Hz). During tooth brushing, controls displayed greater spectral power in the 0-5 Hz range, with three distinct peaks, while PwPD showed only one peak around 4-5 Hz (**Fig. 3c**). This suggests that although both groups share a similar primary movement frequency, PwPD have more diffuse and attenuated frequency contributions outside this range, possibly reflecting difficulty in generating coordinated rotational motions. During hair combing, controls again exhibited greater spectral power in the 0-2 Hz band relative to PwPD, which may result from prolonged, rhythmic combing strokes that are harder for PwPD to sustain (**Fig. 3d**).

### The area of continuous wavelet transform complex plots distinguishes PD patients from controls

PwPD experience a phenomenon known as the sequence effect, characterized by progressive slowing of a repetitive movement [44]. To examine how movement frequency contributions evolved during task performance, we generated continuous wavelet transforms (CWT) of accelerometry data for both PwPD and controls during tooth brushing and hair combing (**Fig. 3e—h**). During brushing, CWT plots corroborated PSD findings; while both groups exhibited a similar primary movement frequency, controls displayed stronger contributions at lower frequencies, whereas PwPD showed more diffuse and attenuated contributions at higher frequencies (**Fig. 3e, g, i**). Likewise, during combing, although PwPD and controls shared a dominant movement frequency, PwPD again demonstrated more diffuse and attenuated high-frequency activity (**Fig. 3f, h, j**). Contrary to expectations of gradual decline in task performance across PwPD trials, we did not observe significant changes in frequency over the course of either brushing or combing tasks.

Previous studies have shown that continuous wavelet transform (CWT) representations in the complex plane often form elliptical patterns, and the area of these ellipses has proven effective in discriminating gait patterns between PwPD and controls [37]. The area of the ellipse formed by plotting the CWT of a signal in the complex plane represents the variability and intensity of the signal. We therefore hypothesized that CWT ellipse area would quantify bradykinesia and serve as a valuable feature for classifying PwPD and control movement data from ADLs. During brushing, PwPD showed a non-significant decrease in ellipse area (*M* = 4.5001 ∗ 10^-9^; *Q*1 = 1.2645 ∗ 10^-9^; *Q*3 = 1.4829 ∗ 10^-8^) compared to controls (*M* = 5.2960 ∗ 10^-9^; *Q*1 = 2.4665 ∗ 10^-9^; *Q*3 = 1.6246 ∗ 10^-8^) (**Fig. 3m, o**). During combing, the reduction in ellipse area was statistically significant for PwPD (*M* = 5.0233 ∗ 10^-9^; *Q*1 = 1.5605 ∗ 10^-9^; *Q*3 = 1.2817 ∗ 10^-8^) relative to controls (*M* = 4.7477 ∗ 10^-8^; *Q*1 = 1.5995 ∗ 10^-8^; *Q*3 = 1.0790 ∗ 10^-7^) (**Fig. 3n, p**). These findings suggest that CWT ellipse area is a physiologically relevant metric for assessing movement alterations in PwPD during combing tasks.

### XGBoost classification with short intervals of ADL-based movement data differentiates severity and identifies cardinal motor features in PD

Having identified key differences in ADL-based movement data between PwPD and controls, we hypothesized that these data could be used to train ML models to predict specific clinical outcomes, including the presence of cardinal motor symptoms and overall disease severity. We trained and evaluated XGBoost classification models using 30 features extracted from non-overlapping signal intervals of 3, 5, 7, and 9 seconds. For each task, model performance was measured by averaging balanced accuracies (BA) across all cross-validation (CV) folds and constructing a 95% confidence interval using the ten repeats (**Fig. 4**). We show this average (BA) across all classification tasks (**A1-A7 and B1; Supplemental Table 3)**.

**Figure 4.**
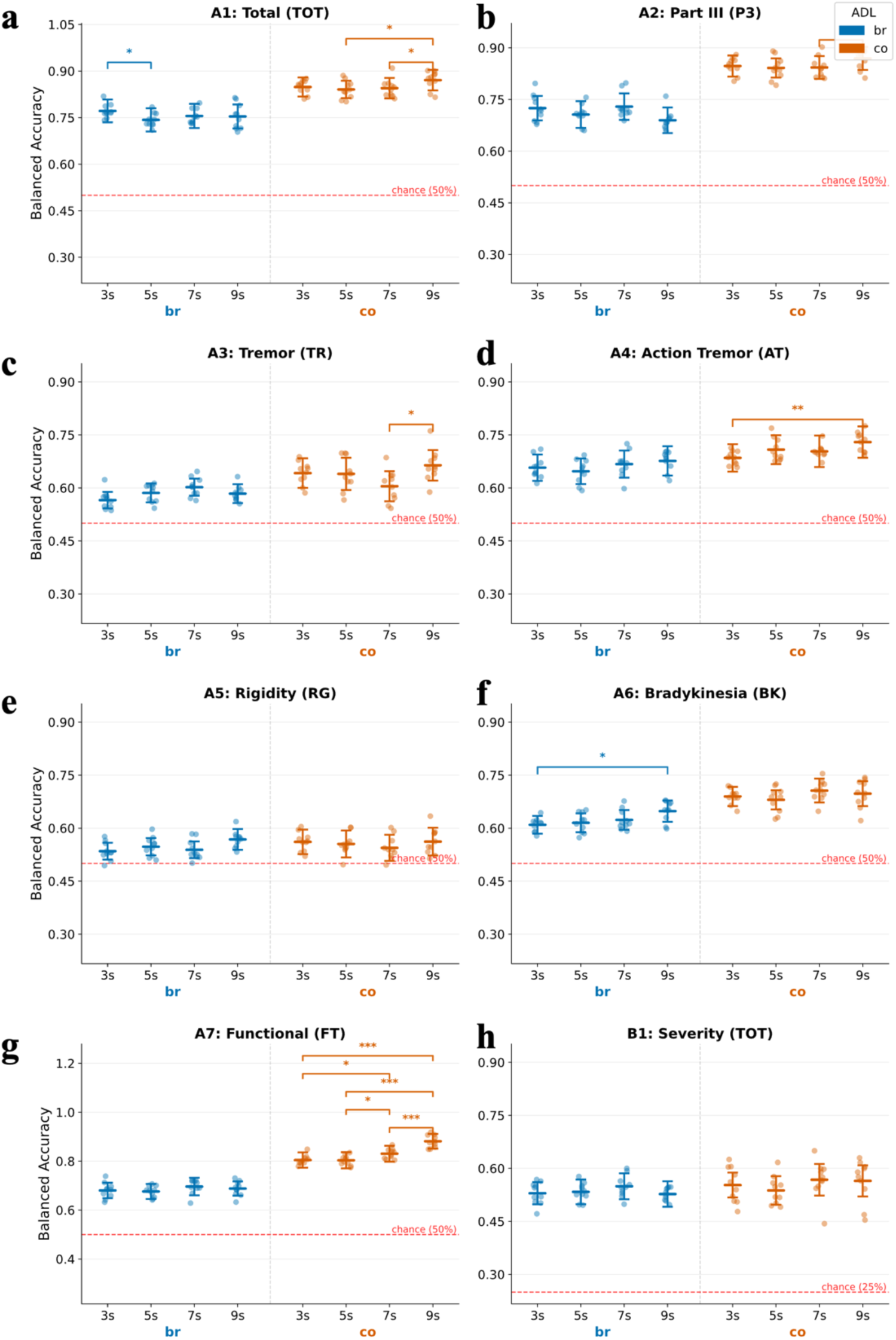
Cross-validation results for XGBoost classification tasks A1-A7 and B1 with pairwise comparisons across interval lengths. Each point corresponds to a mean balanced accuracy (BA) across all (10) folds within one repeat. Brushing results are colored in blue, and combing tasks are colored in orange, plotted above a red-dotted line indicating chance-level performance for the task. Bars correspond to 95% confidence intervals about the mean. Paired t-tests with Holm-Bonferroni correction were used to assess significance (*p < 0.05, **p < 0.01, ***p < 0.001). Extreme outliers were 95% winsorized.

Models trained on both toothbrushing and hair combing data consistently outperformed chance across all fold lengths and tasks (**Table 2**). Notably, prediction performance was strongest for motor scores (P3), action tremor (AT), bradykinesia (BK), and functionality (FT), and weakest for rest tremor (TR) and rigidity (RG) (**Fig. 4b-g**). This pattern likely reflects the differential expression of these symptoms during ADLs; for instance, action tremor would be expected to manifest more prominently during ADL performance than rest tremor.

**Table 2.** Cross-validation balanced accuracy (BA) scores across tasks and interval lengths. Mean BA **±** standard deviation across ten repeats of ten-fold cross validation is presented for each task and interval length. Longer interval lengths tended to yield higher BA.

|  | Task | 3s | 5s | 7s | 9s |
|---|---|---|---|---|---|
| brush | A1 | **0.770 ± 0.188** | 0.742 ± 0.191 | 0.754 ± 0.199 | 0.752 ± 0.197 |
|  | A2 | 0.723 ± 0.182 | 0.705 ± 0.199 | **0.726 ± 0.198** | 0.690 ± 0.190 |
|  | A3 | 0.564 ± 0.133 | 0.585 ± 0.150 | **0.602 ± 0.133** | 0.583 ± 0.145 |
|  | A4 | 0.653 ± 0.198 | 0.645 ± 0.187 | 0.663 ± 0.204 | **0.669 ± 0.225** |
|  | A5 | 0.534 ± 0.126 | 0.546 ± 0.132 | 0.537 ± 0.128 | **0.567 ± 0.157** |
|  | A6 | 0.608 ± 0.135 | 0.614 ± 0.145 | 0.619 ± 0.152 | **0.647 ± 0.164** |
|  | A7 | 0.674 ± 0.176 | 0.676 ± 0.161 | **0.695 ± 0.185** | 0.686 ± 0.152 |
|  | B1 | 0.528 ± 0.167 | 0.534 ± 0.185 | **0.547 ± 0.198** | 0.523 ± 0.195 |
| comb | A1 | 0.835 ± 0.188 | 0.834 ± 0.156 | 0.834 ± 0.199 | **0.864 ± 0.187** |
|  | A2 | 0.834 ± 0.188 | 0.835 ± 0.157 | 0.831 ± 0.200 | **0.863 ± 0.189** |
|  | A3 | 0.636 ± 0.224 | 0.629 ± 0.250 | 0.600 ± 0.228 | **0.655 ± 0.234** |
|  | A4 | 0.669 ± 0.230 | 0.700 ± 0.223 | 0.695 ± 0.242 | **0.717 ± 0.250** |
|  | A5 | 0.559 ± 0.183 | 0.554 ± 0.202 | 0.546 ± 0.192 | **0.561 ± 0.210** |
|  | A6 | 0.687 ± 0.148 | 0.676 ± 0.152 | **0.703 ± 0.178** | 0.697 ± 0.184 |
|  | A7 | 0.794 ± 0.191 | 0.792 ± 0.200 | 0.817 ± 0.204 | **0.869 ± 0.193** |
|  | B1 | 0.552 ± 0.188 | 0.536 ± 0.217 | **0.567 ± 0.235** | 0.563 ± 0.235 |

Models trained on hair combing data outperformed those trained on toothbrushing data across all tasks except RG (**Fig. 4e**). We found no consistent relationship between interval length and CV performance. For most tasks, no interval length significantly outperformed others, with the exception that nine-second intervals yielded significantly better performance than three-second intervals. For combing models trained to predict FT, both seven- and nine-second intervals significantly outperformed three- and five-second intervals (**Fig. 4g**).

### Feature importance analysis reveals distinct sensor features drive predictions for brushing versus combing

To understand which features drove our models’ predictions, we generated volcano plots to compare the effect size and significance between PwPD and controls for each extracted feature. For both tasks, acceleration-derived features generally exhibited larger effect sizes, whereas angular-velocity-derived features reached higher significance levels, indicating that acceleration features may be more variable between groups, but less discriminative than angular velocity features (**Fig. 5a, b**).

**Figure 5.**
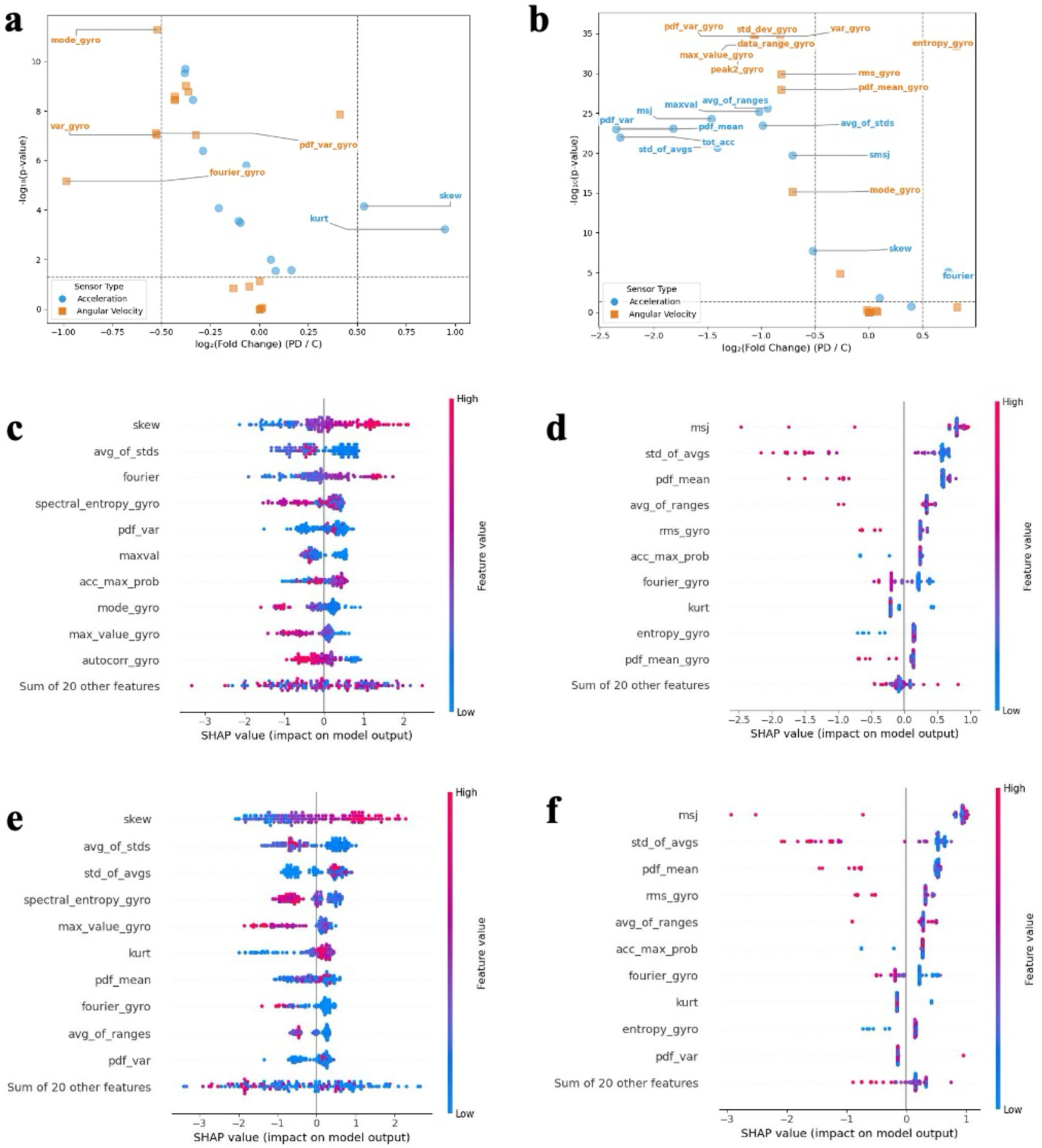
Identification of key features predictive of motor symptom severity in PD. Feature importance from acceleration and angular velocity data during toothbrushing (left panels) and hair-combing (right panels) is analyzed using two complementary methods. (a-b) Volcano plots showing the significance versus effect size of features from PwPD compared to controls. (c-f) SHAP summary (beeswarm) plots depicting the top 10 most predictive features for regression models estimating clinical severity: the UPDRS motor Part III (P3; c-d, brush, comb) and summed Part I-III scores (TOT; e-f, brush, comb).

From brushing data, only four angular velocity-based features and two acceleration-based features surpassed the effect-size threshold (+/-0.5 log2 Fold Change). Both acceleration features had a positive effect, while all four angular velocity features had a negative effect. From combing data, in contrast, a more features from each sensor type exceeded the effect-size threshold (ten angular-velocity features, eleven acceleration features), but angular-velocity features were markedly more significant. This indicates that angular velocity may be particularly discriminative when modeling combing movements. The most significant feature for brushing was the mode of the angular velocity distribution, and, for combing, the standard deviation & variance of the angular velocity distribution.

We additionally used SHAP analysis to corroborate our findings. We generated beeswarm plots to visualize feature importance for the XGBoost models predicting P3 (**Fig. 5c, d**) and TOT scores (**Fig. 5e, f**). These plots showed that the top predictive features for brushing were consistently dominant frequency and skew of the acceleration distribution; as well as spectral entropy of the angular velocity distribution. Additionally, certain window metrics measuring intra-signal variability (“std_of_avgs”) and overall spread (“avg_of_stds”) were important. In combing models, however, the most important features for both P3 and TOT predictions were consistently mean squared jerk and mean of the acceleration distribution, along with window metrics. Acceleration-based features seemed to have a higher effect on model classification.

## IV. Discussion

Our preliminary findings establish the utility of ADL-based movement data for assessing PD symptoms and severity, with several potential applications for the direct evaluation of FA and QOL in PwPD. We generated KDE plots to assess distributional trends in acceleration and angular velocity data collected from PwPD of varying disease severity and controls. We found that patients with increased severity tended to have narrower distributions near the origin, which we attribute to more pronounced bradykinesia and corresponding movement decrements. We corroborated our KDE analyses by comparing signal energy; while combing their hair, PwPD had less signal energy than controls.

Using PSD and CWT analyses, we found that while both populations shared a dominant frequency across tasks, controls tended to have greater low-frequency contribution, whereas PwPD had greater, diffuse high-frequency content. Surprisingly, we did not observe the sequence effect upon CWT analysis.

This may be attributable to patient overperformance of goal-oriented tasks under observation in clinic or the short 30s duration of our analyses. Building on previous findings, we identified a statistically significant difference in CWT complex plots areas between PwPD and controls while combing, which we believe is a valuable feature for future investigations.

Furthermore, we determined that XGBoost models trained on very short intervals (as low as three seconds) of brushing and combing data can feasibly identify PwPD, functional impairment, and even some cardinal symptoms. Changing input interval length did not consistently impact performance, despite our expectation that longer input intervals would yield better models. While this was true for some tasks, the relationship did not hold across ADLs within a given task, and across tasks. We note that A3 (rest tremor) and A5 (rigidity) performed the most poorly amongst all aggregate scores calculated, with balanced accuracies only slightly above chance. This mirrored our expectations; rest tremors should not be detectable during ADL performance, and rigidity is typically measured with a limp arm guided by the clinician rather than with task performance. In contrast, A7 (functional score) which is more directly relevant to ADL performance was predicted with much higher BA.

To understand which features drove model predictions, we performed SHAP analysis and generated volcano plots to identify the top ten most predictive features. Acceleration-based features seemed have the largest effect on model classification, with simple distributional features like skew, mean squared jerk, dominant frequency, and variance labeled as the highest importance features.

Our study has several important limitations. First, this is a pilot analysis with a small sample, notably of healthy controls (n = 7, compared to n = 39 PwPD). Class imbalance may limit the stability of our performance estimates. Study enrollment is ongoing, and we plan to recruit additional healthy controls and PwPD. Second, the enrolled PwPD cohort was only collected from within the Johns Hopkins system and lacked heterogeneity in disease severity, with a majority (76.9%) classified as mild (TOT: 1-32). Only those who could perform the toothbrushing and combing tasks without excessive difficulty or pain participated in the study. This limits the generalizability of our findings to individuals with moderate or advanced PD. We also note some participants experienced upper-extremity dyskinesias during their participation, as measured with the Abnormal Involuntary Movement Scale (AIMS) (n = 3/33; six PwPD could not be evaluated) [43]. These participants generally produced acceleration distributions like those of their severity-matched cohorts, but they also yielded angular velocity distributions more concentrated around the origin **(Supplemental Fig. 1).** Relatedly, medication status (ON/OFF) was not tracked, which may misrepresent functional ability; participants that were OFF may underperform greatly. Third, while we attempted to age-match controls by recruiting spouses and caregivers of PwPD, there remains some inherent variability within our control group. Some participants were younger, and elderly controls may have exhibited age-related motor impairments, meaning their assumed UPDRS score of zero may be too reductionistic. This could reduce the contrast between patient and control groups. Finally, the 30-second duration of our task analyses and the clinic-based observation setting may have influenced participant behavior (e.g., overperformance), potentially masking symptoms like decrement of amplitude (the sequence effect) [44].

Compared to existing sensor-based wearables and predictive systems, our approach demonstrates significant potential for providing context-specific, task-based assessments that directly reflect real-world functional ability. Many continuous monitoring systems generate a vast quantity of clinical data, much of which is useful for characterization of patient disease state; however, much of it is non-contextualized, requiring separate interpretation and integration into time-constrained clinical workflows [45]. In contrast, our method addresses this challenge by requiring only minutes of targeted, task-specific data collection, thereby enabling more focused, interpretable, and clinically actionable assessments.

Our study also contributes a valuable dataset of signals recorded during ADLs from PwPD and controls. This dataset represents a meaningful improvement over resources currently available in the literature. Existing datasets are often limited by one or more of the following: 1) inclusion of only healthy controls; 2) very small sample sizes per experimental group (*n* ≤ 10); 3) reliance on experimental, non-standardized tools (e.g., self-stabilizing prototypes); or 4) limited contextual information for the recorded signals [28, 46].

One recent study reported a machine learning model for detecting wrist tremor and akinesia with accuracies of 97% and 100%, respectively, validated against MDS-UPDRS scores [47]. However, this model was trained on data from only 10 patients. In contrast, our best models across interval lengths— trained on data from a substantially larger cohort— achieved BAs of 0.669 (action tremor) using brushing data, and 0.717 using combing data. Notably, our model predicted bradykinesia from combing data with a BA of 0.703. By employing cross-validation, our results demonstrate robust performance on this larger dataset of participants.

We posit that our method could be adapted to other instrumented ADLs, such as utensil use, and potentially to more complex, unstructured tasks like dressing. Furthermore, the need for objective functional assessment extends beyond PD to other movement disorders, including atypical parkinsonisms, Essential Tremor, and limb-based dystonias. These conditions are amenable to the same analytical pipeline—kernel density estimation, continuous wavelet transforms, and feature extraction for machine learning—to derive disorder-specific motor signatures. Further validation is also needed to investigate whether our approach could feasibly track task performance longitudinally.

In summary, this work establishes a promising framework for objective monitoring of functional motor ability. By translating brief, context-specific tasks into quantifiable metrics, this approach can propel the movement disorders field closer to personalized, patient-centered care and deepen our understanding of how neurodegenerative disease impacts daily life.

## Data Availability

Patient-level data used in this study were derived from electronic health records and cannot be shared due to restrictions under the IRB-approved protocol. Aggregate results supporting the conclusions are contained in the manuscript.

## V. Acknowledgements

The authors would like to thank the Johns Hopkins University Department of Biomedical Engineering, particularly Michelle Zwernemann, MSE, for providing discretionary funds and mentorship. Lastly, we would like to thank Ryan Roemmich, PhD, and James Brasic, MD, MPH, for their long-term mentorship and guidance.

