## Supplemental Materials for "Short intervals of inertial measurement unit data collected during activities of daily living can provide a quantitative method to assess disease severity and functional ability in Parkinson’s disease"

- **Supplemental Table 1.** UPDRS composite score definitions
- **Supplemental Table 2.** Signal feature definitions
- **Supplemental Table 3.** Classification Testing Scheme
- **Supplemental Figure 1.** Kernel density estimation plots for positive upper extremity score on Abnormal Involuntary Movement Scale

| Aggregate Score | Subscores (Part.Number) used in calculation |
|---|---|
| Rigidity (RG) | 3.22 Rigidity (neck), 3.22 Rigidity (right arm) OR (left arm) |
| Bradykinesia (BK) | 3.23 Finger Taps (right) OR (left), 3.24 Hand Movements (right) OR (left) 3.25 Rapid alternating movements of hand (right) OR (left) |
| Functional ADL (FT) | 2.6 Salivation, 2.7 Swallowing, 2.8 Handwriting, 2.9 Cutting food and handling utensils, 2.10 Dressing, 2.11 Hygiene |
| Resting Tremor (TR) | 3.20 Tremor at rest (head), 3.20 Tremor at rest (right arm), 3.20 Tremor at rest (left arm), 3.20 Tremor at rest (right leg), 3.20 Tremor at rest (left leg) |
| Action/Postural Tremor (AT) | 3.21 Action or Postural Tremor of Hands (right) OR (left) |
| Part III Sum (P3) | All Part III UPDRS subscores |
| Total Sum (TOT) | All Part I-III UPDRS subscores |

**Supplementary Table 1**. UPDRS subscores comprising the calculated aggregate scores. These scores quantify distinct motor dimensions of patient symptomatology and serve as target variables in classification tasks. Except for resting tremor, which reflects the whole-body rather than hand used during the activity, all subscores correspond to the hand used during participation.

| Feature | Variable Name | Definition |
|---|---|---|
| Skewness | skew | Skewness is a statistical measure of the asymmetry of a probability distribution of acceleration data |
| Kurtosis | kurt | The degree to which distribution of acceleration data is concentrated in the center or in the tails of the distribution, relative to a normal distribution |
| Mean Squared Jerk | msj | The average of the squared first derivative of the acceleration signal |
| Scaled Mean Squared Jerk | smsj | The mean squared jerk scaled by the largest absolute value jerk value in the signal. |
| Maximum Value | maxval | The maximum value of the acceleration signal |
| Mean | pdf_mean | The mean value of the acceleration distribution |
| Variance of the Distribution | pdf_var | Variance of the acceleration distribution |
| Mode of Acceleration Distribution | acc_max_prob | The value of the mode of the acceleration distribution |
| Total Acceleration | tot_acc | Sum of the average acceleration in each sliding window of the signal [48] |
| Average of Window Ranges | avg_of_ranges | Average of range of acceleration values in each sliding window of the signal |
| Average of Window Standard Deviations | avg_of_std | Average of standard deviation of acceleration values in each sliding window of the signal |
| Standard Deviation of Window Means | std_of_avgs | Standard deviation of the mean of acceleration values for each sliding window of the signal |
| Dominant Frequency | fourier | The position of the dominant frequency within the produced array of frequencies of the acceleration signal |
| Skewness | skew_gyro | Skewness is a statistical measure of the asymmetry of a probability distribution of angular velocity data |
| Kurtosis | kurt_gyro | The degree to which distribution of angular velocity data is concentrated in the center or in the tails of the distribution, relative to a normal distribution |
| Maximum Value | max_value_gyro | The maximum value of angular velocity signal |
| Peak Value | gyro_max_peak | The value of the KDE distribution at the mode |
| Fourier Transform | fourier_gyro | The calculated magnitude spectrum of frequencies within the angular velocity signal |
| Mean | pdf_mean_gyro | Mean of the angular velocity distribution |
| Variance of the Distribution | pdf_var_gyro | Variance of the angular velocity distribution |
| Mode | mode_gyro | The mode of the distribution of angular velocity |
| Standard Deviation | std_dev_gyro | The standard deviation of the signal of angular velocity values |
| Variance | var_gryo | The variance of the signal of angular velocity values |
| Range | data_range_gyro | The difference between the minimum and maximum of |

| | | |
|---|---|---|
| | | the signal of angular velocity values |
| Root Mean Square | rms_gyro | The magnitude of a varying quantity, calculated as the square root of the arithmetic mean of the squares of a set of angular velocity values. |
| Peak to Peak Difference | peak2_gyro | The difference between the highest (peak) and lowest (trough) values of the angular velocity signal |
| Autocorrelation | autocorr_gyro | The similarity between the angular velocity signal and a delayed copy of itself as a function of the delay. A measure of periodicity within the angular velocity signal |
| Zero Crossing Rate | zcr_gyro | The number of times a signal crosses the zero-amplitude line within a given time interval |
| Entropy | entropy_gyro | The measure of the uncertainty or randomness of the angular velocity signal |
| Spectral Entropy | spectral_entropy_gyro | The randomness or unpredictability in the frequency distribution of signal |

**Supplementary Table 2**. Definitions for 30 features extracted from each signal interval. Alongside descriptive statistics, features characterize the signal distribution, spectral representation, temporal representation, and peak characteristics. Features were handpicked after inspecting both the original time-series signals and KDE plots, as well as reviewing current approaches in the literature.

| | TOT | P3 | TR | AT | RG | BK | FT |
|---|---|---|---|---|---|---|---|
| A. Identification | A1 | A2 | A3 | A4 | A5 | A6 | A7 |
| B. Severity | B1 | | | | | | |

**Supplementary Table 3**. All classification tasks performed (A1-A7, B1) for all interval lengths (3s, 5s, 7s, 9s). Tasks in row A indicate binary classification tasks in which the decision tree separates input data into classes of zero and nonzero value of the target variable (in the columns). Row B indicates classification tasks with multiple bins based on their UPDRS Part I-III aggregate score.

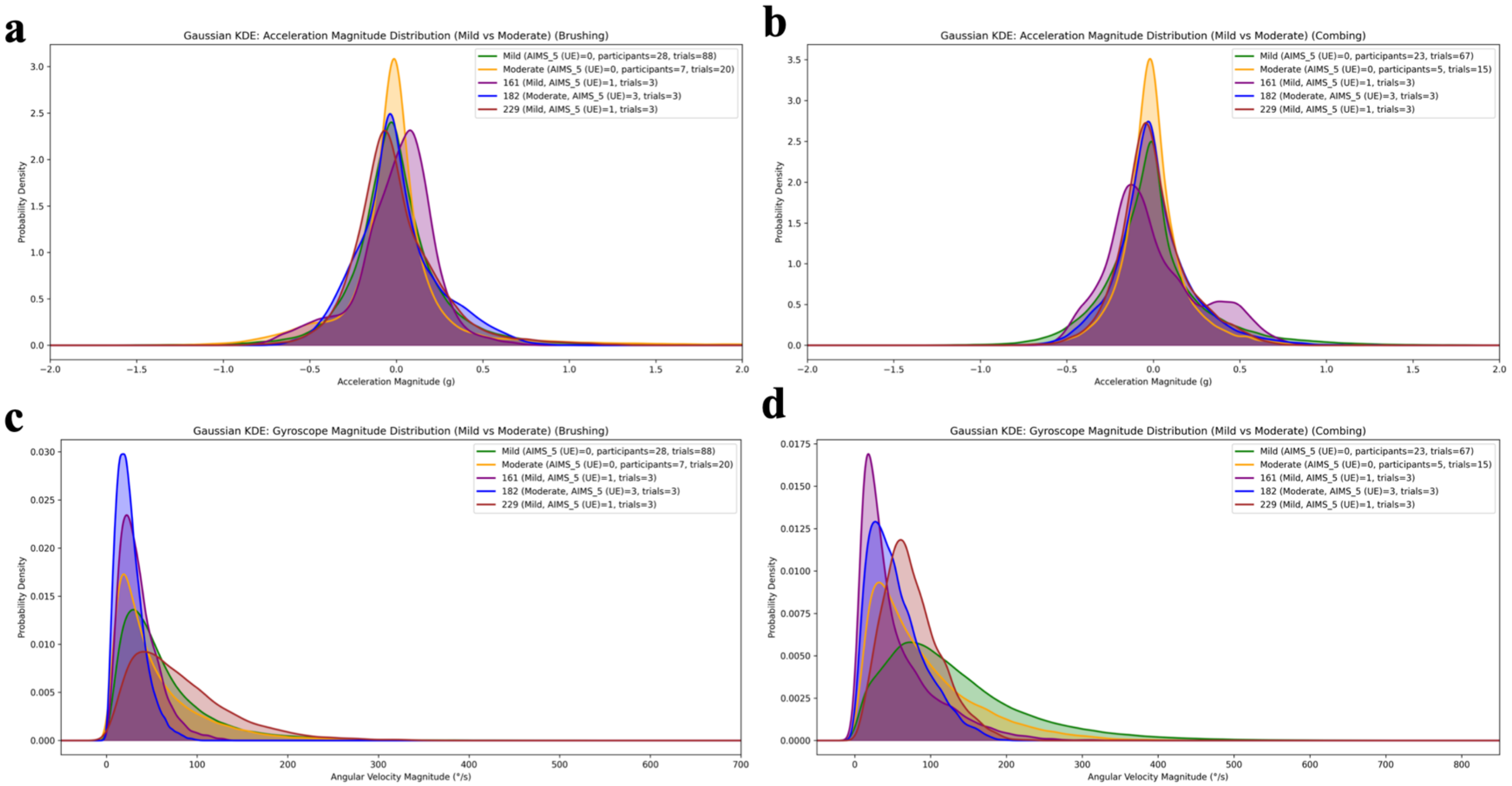


**Supplemental Figure 1:** Kernel density estimation (KDE) plots showing three participants with positive AIMS upper extremity score compared to rest of the cohort. Left panels show brushing performance and right panels show combing performance. (a,b) Acceleration distribution during the brushing and combing task for three positive participants compared to all mild and moderate participants. (c,d) Angular velocity distribution during the brushing and combing task for the same participants.
